# Demographic and Hygienic Factors as Predictors of Face Mask Wearing During Covid-19 Pandemic in Malaysia

**DOI:** 10.1101/2021.02.09.21251280

**Authors:** Kim Hoe Looi, Stephen X. Zhang, Nicolas Li

**Affiliations:** Xiamen University Malaysia, Malaysia; University of Adelaide, Australia; University of Dundee, United Kingdom

**Keywords:** Covid-19, face mask wearing compliance, demographic factors, hygienic factors, Malaysia

## Abstract

Wearing a face mask has been recognised as an effective way of slowing down the spread of the Covid-19 pandemic. However, there is scarce evidence on predictors of face mask wearing during a pandemic. This research aims to investigate which demographic and hygienic factors could predict the compliance for face mask wearing in Malaysia. We employed a structured online survey of 708 Malaysian adult respondents. Among the factors examined, we found gender, hand washing and wearing of personal protective equipment significantly predicted face mask wearing.

## 1. Introduction

It has been over six months since the World Health Organisation (WHO) declared Covid-19 a pandemic. As the number of infections kept increasing at an alarming rate, various actions had been taken to reduce transmission of Covid-19. The primary non-pharmaceutical interventions at the population level include border closure and public places shutdown. At the individual level, the primary non-pharmaceutical interventions include wearing face mask, improved hygiene (e.g., washing hands) and other physical barriers (e.g., physical distancing), until an effective and cost-efficient vaccine becomes widely available (i.e., pharmaceutical intervention).

Health agencies worldwide recommended wide compliance of wearing face mask in public settings during the Covid-19 pandemic. For example, the US Centers for Disease Control and Prevention (CDC) recommended: “Cover your mouth and nose with a mask when around others” [1]. WHO recommended use of personal protective equipment (PPE) – such as medical/surgical face masks – as part of a comprehensive strategy of infection prevention and control (IPC) against transmission of COVID-19 [2, 3]. Although face mask wearing by the healthy population to reduce risk of transmission of Covid-19 remains controversial [4,5], an increasing number of latest literature are recommending community-wide face mask wearing [3, 4, 6-9]. For instance, modelling results suggest a potentially high value of wearing face mask by the general public to curtail community transmission of Covid-19 [5]. It is advocated that the precautionary principle on the grounds that there is little to lose and potentially something to gain from wearing face mask [10].

Therefore, before the arrival of vaccine, governments should encourage the general population in specific situations and public settings to wear face masks to protect healthy persons (i.e., prevention) and suppress onward transmission of Covid-19 by an infected individual to others (i.e., source control) [3-5]. Face mask must be used by all people most of the time and in a correct manner to be effective [10]. Nevertheless, wearing face mask per se is insufficient to prevent infection of Covid-19 [3, 8] and may create a false sense of safety [10]. Community-wide adoption and high compliance of wearing face mask in conjunction with other non-pharmaceutical interventions (for example, hand hygiene and physical distancing) are critical to prevent human-to-human transmission of COVID-19 [3, 5, 8].

Wearing a face mask proffers various benefits, as it is relatively cheap, simple operation, strong sustainability, high health benefits and good health economy [5, 6, 8-11]. However, the effectiveness of face masks for prevention and source control depends on individuals’ compliance [11]. It was noted a lower level of face masks compliance or lower reported acceptability vis-à-vis hand hygiene and other non-pharmaceutical interventions [11]. To date, very few studies seek to measure levels of face mask compliance or predict the level of face mask compliance during Covid-19 pandemic. Behavioural compliance depends on many issues, ranging from availability, cost, discomfort and breathing difficulty from prolonged face mask wearing, demographics, to local contexts. Scholars have noticed different receptiveness to face mask wearing across countries [9]. For example, in some east Asian countries, face mask wearing is ubiquitous and mandatory [6-8, 10]. In certain countries, many people are reluctant and/or oppose to wear face masks, regarding it as a symbolic choice of freedom [6-8].

The Malaysian Ministry of Health has been advocating wearing face mask as a complementary personal protective device against Covid-19 and has tirelessly conducted many health campaigns to prevent Covid-19, with face mask wearing an essential element [12]. Face mask wearing in crowded public places is mandatory in Malaysia from 1st August 2020, with a fine of RM1,000 (approximately USD240) for violation. As positive cases of Covid-19 continue to shoot up, on 3rd October 2020, the Director General of Ministry of Health reminded that “public health measures can also be carried out by each and every individual, by practising the three Ws and avoiding the three Cs,” that is, to always wash hands, wear a face mask and remember warnings by the authorities as well as to avoid confined spaces, crowded places and close quarters conversations [13].

Wearing a face mask in public places is a new normal and has become the global symbol to fight Covid-19 pandemic. This study investigates levels of face mask wearing (compliance) and explains the levels of face mask compliance during Covid-19 pandemic in Malaysia employing demographic factors and hygienic factors.

## 2. Methods

We conducted our investigation using a web-based cross-sectional questionnaire in Malaysia, which was a safe and feasible way of collecting data during the pandemic, in line with other similar studies [14].

Given the multicultural nature of Malaysia, the questionnaire is available in three major languages (i.e., Malay, Mandarin and English) and links to the questionnaire were distributed via WhatsApp, Facebook Messenger and email. To minimise response and measurement biases, we followed the standard survey approaches [15], that is, no social pressure to influence responses, no questions that provoke defensiveness or threaten esteem, no payoff or cost for particular responses. Multi-item questions were used to ensure no priming, and there was no overlapping among questions for different constructs [16]. Participation in this survey was voluntary, and respondents could opt out at any time. Moreover, respondents were assured anonymity and confidentiality of their responses. All respondents consented online before proceeding to answer the questionnaire.

Data collected include respondents’ demographic characteristics (gender, age, educational level, and the number of children under 18 years-old living in the same household) and selfreported hygienic measures (hand washing and wearing of PPE).

### 2.1 Gender

The variable gender was a dichotomous variable coded 1 if the respondent was male and 2 if female.

### 2.2 Age

Respondents reported their year of birth. Their age ranged from 21 years-old to 71 years-old.

### 2.3 Number of children in the household

This variable was measured from 1 (none) to 7 (six or more children).

### 2.4 Handwashing

Handwashing is one of the most basic ways of personal hygiene [17]. This variable was measured from 1 (never) to 7 (every time). Pearson correlation shows a medium positive strength between handwashing and face mask wearing (0.29 at the 0.01 level) and attests to a low probability of multicollinearity.

### 2.5 Wearing of PPE

We asked the respondents if they have sufficient personal protective equipment, measured from 1 (never) to 5 (always). Pearson correlation shows a medium positive strength between PPE and face mask wearing (0.45 at the 0.01 level).

### 2.6 Face mask wearing

We asked the respondents how frequent they wear a facemask when outside of their houses. The response scale ranges from 1 (never) to 7 (every time).

We analysed the data using SPSS (v. 26) and used multivariate least-squares fitting analysis to predict the behaviour of wearing a face mask at the significance level of 0.05.

## 3. Results

### 3.1. Descriptive Findings

We received 708 valid responses (data available upon request). Table 1 shows the descriptive statistics of the sample. The percentage of male and female respondents was almost equivalent. The sample was also represented by diverse age groups and the number of children in the household. In contrary to Cowling et al. [11], this study found a higher level of face mask compliance vis-à-vis hand hygiene. Face mask wearing is very visible, hence, higher compliance in a collectivistic society such as Malaysia, relative to hand washing, which is less visible to surrounding people. (Note: Data for this study was collected prior to mandatory face mask wearing in public places)

**Table 1.**
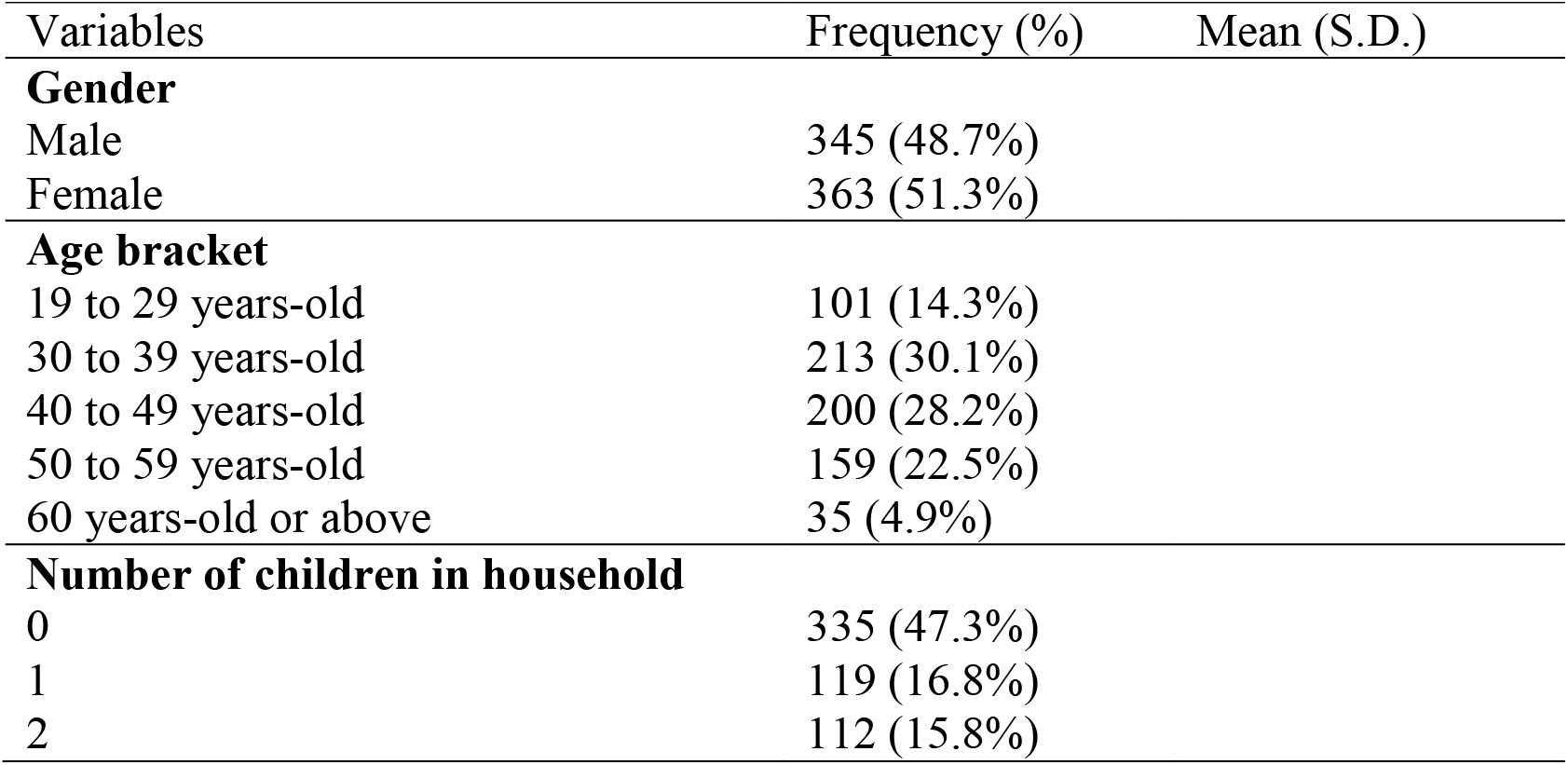
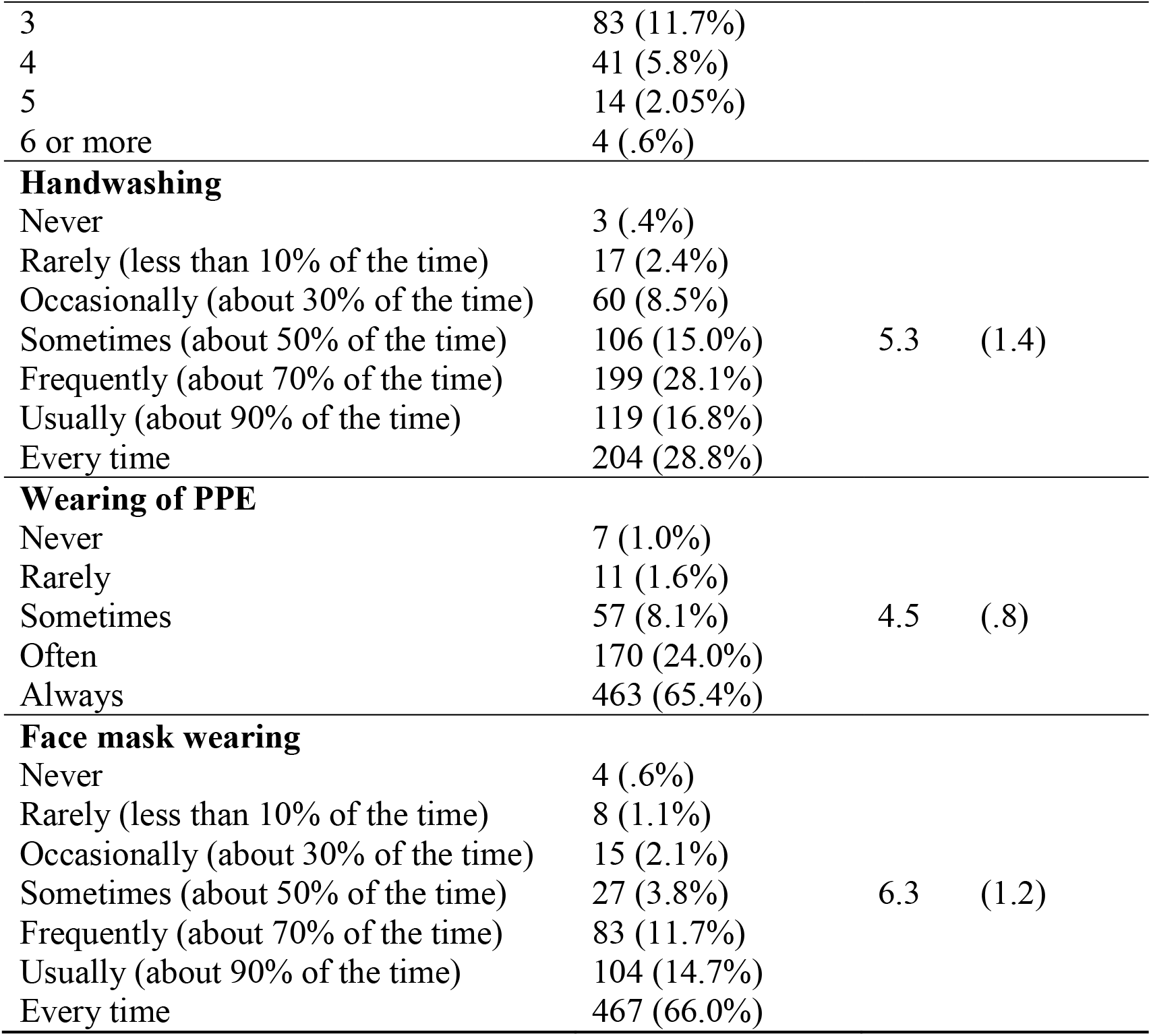
Descriptive statistics of respondents

| Variables | Frequency (%) | Mean (S.D.) |
| --- | --- | --- |
| **Gender** |  |  |
| Male | 345 (48.7%) |  |
| Female | 363 (51.3%) |  |
| **Age bracket** |  |  |
| 19 to 29 years-old | 101 (14.3%) |  |
| 30 to 39 years-old | 213 (30.1%) |  |
| 40 to 49 years-old | 200 (28.2%) |  |
| 50 to 59 years-old | 159 (22.5%) |  |
| 60 years-old or above | 35 (4.9%) |  |
| **Number of children in household** |  |  |
| 0 | 335 (47.3%) |  |
| 1 | 119 (16.8%) |  |
| 2 | 112 (15.8%) |  |

|  |  |  |  |
| --- | --- | --- | --- |
| 3 | 83 (11.7%) |  |  |
| 4 | 41 (5.8%) |  |  |
| 5 | 14 (2.05%) |  |  |
| 6 or more | 4 (.6%) |  |  |
| **Handwashing** |  |  |  |
| Never | 3 (.4%) |  |  |
| Rarely (less than 10% of the time) | 17 (2.4%) |  |  |
| Occasionally (about 30% of the time) | 60 (8.5%) |  |  |
| Sometimes (about 50% of the time) | 106 (15.0%) | 5.3 | (1.4) |
| Frequently (about 70% of the time) | 199 (28.1%) |  |  |
| Usually (about 90% of the time) | 119 (16.8%) |  |  |
| Every time | 204 (28.8%) |  |  |
| **Wearing of PPE** |  |  |  |
| Never | 7 (1.0%) |  |  |
| Rarely | 11 (1.6%) |  |  |
| Sometimes | 57 (8.1%) | 4.5 | (.8) |
| Often | 170 (24.0%) |  |  |
| Always | 463 (65.4%) |  |  |
| **Face mask wearing** |  |  |  |
| Never | 4 (.6%) |  |  |
| Rarely (less than 10% of the time) | 8 (1.1%) |  |  |
| Occasionally (about 30% of the time) | 15 (2.1%) |  |  |
| Sometimes (about 50% of the time) | 27 (3.8%) | 6.3 | (1.2) |
| Frequently (about 70% of the time) | 83 (11.7%) |  |  |
| Usually (about 90% of the time) | 104 (14.7%) |  |  |
| Every time | 467 (66.0%) |  |  |

### 3.2. Inference findings

Table 2 summarises the analysis of variance (ANOVA) results. There is a statistically significant difference in face mask wearing between male and female.

**Table 2.**
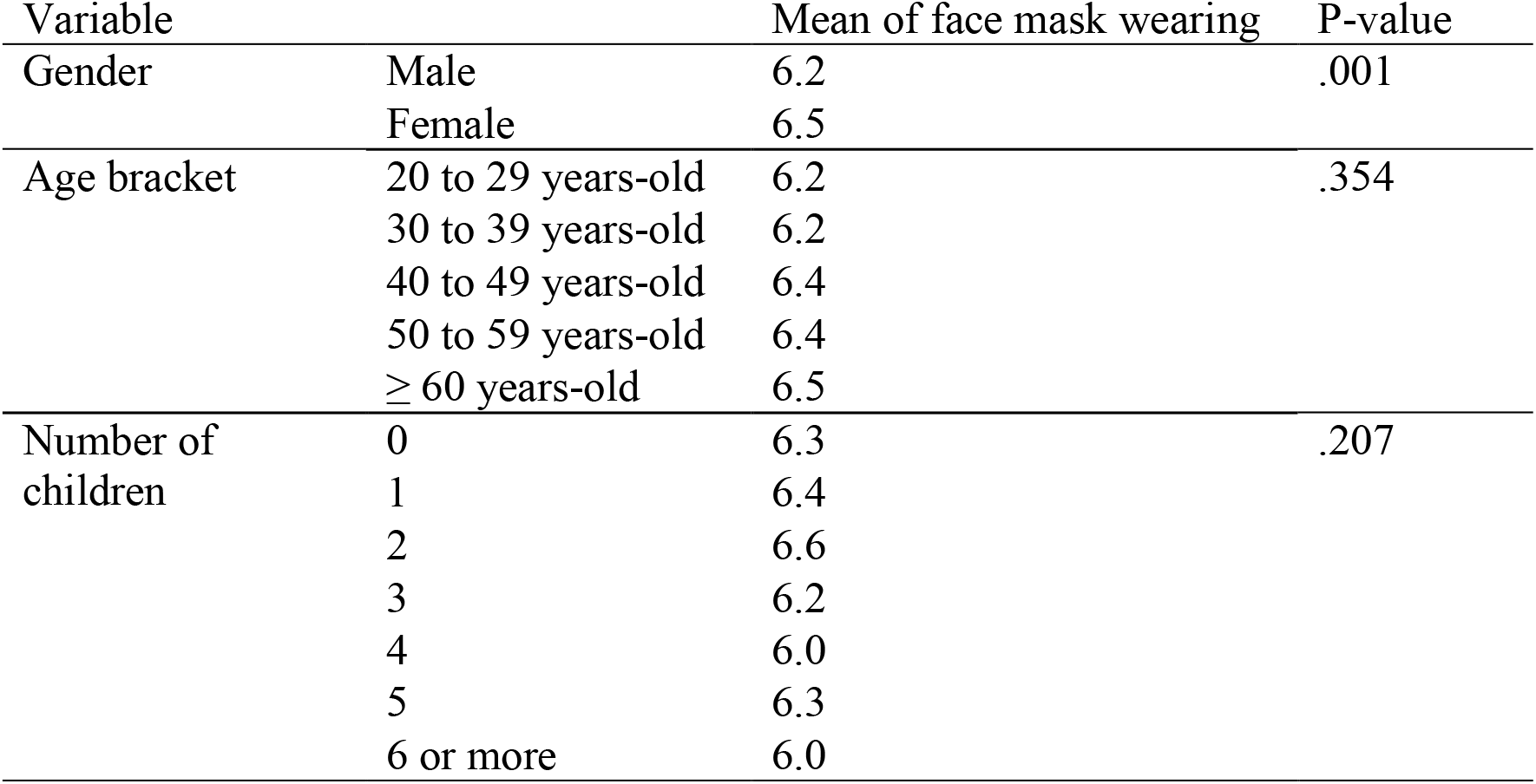
Summary of ANOVA

| Variable |  | Mean of face mask wearing | P-value |
| --- | --- | --- | --- |
| Gender | Male | 6.2 | .001 |
|  | Female | 6.5 |  |
| Age bracket | 20 to 29 years-old | 6.2 | .354 |
|  | 30 to 39 years-old | 6.2 |  |
|  | 40 to 49 years-old | 6.4 |  |
|  | 50 to 59 years-old | 6.4 |  |
|  | ≥ 60 years-old | 6.5 |  |
| Number of children | 0 | 6.3 | .207 |
|  | 1 | 6.4 |  |
|  | 2 | 6.6 |  |
|  | 3 | 6.2 |  |
|  | 4 | 6.0 |  |
|  | 5 | 6.3 |  |
|  | 6 or more | 6.0 |  |

Table 3 presents multiple regression to predict face mask wearing. Among demographic factors, gender positively predicted face mask wearing (p < 0.05) but not age and number of children in the household. In terms of personal hygiene factors, both handwashing and wearing of PPE were strong and positive predictors of face mask wearing (p < 0.01).

**Table 3.**
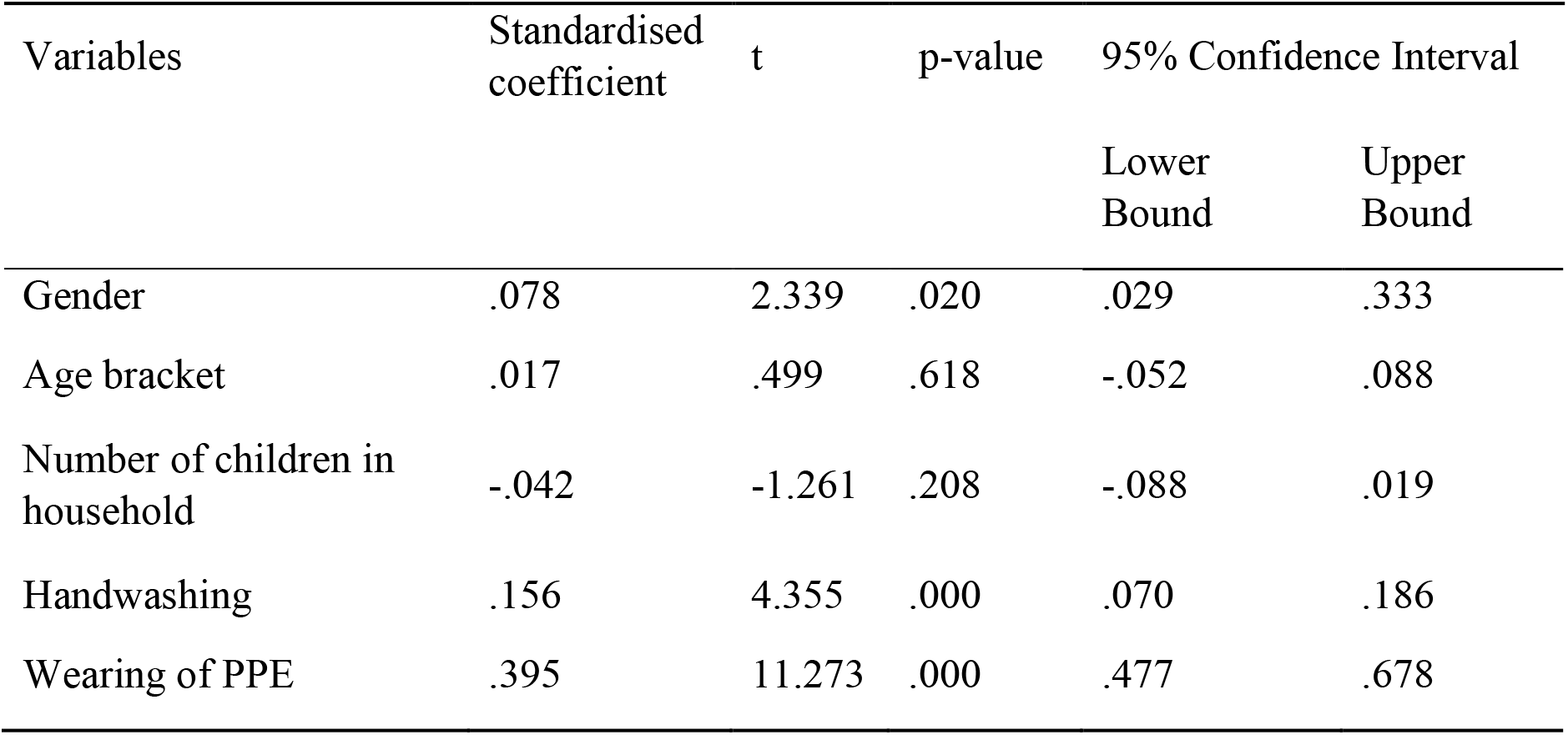
Predictors of face mask wearing (n = 708)

| Variables | Standardised coefficient | t | p-value | 95% Confidence Interval |  |
| --- | --- | --- | --- | --- | --- |
|  |  |  |  | Lower Bound | Upper Bound |
| Gender | .078 | 2.339 | .020 | .029 | .333 |
| Age bracket | .017 | .499 | .618 | -.052 | .088 |
| Number of children in household | -.042 | -1.261 | .208 | -.088 | .019 |
| Handwashing | .156 | 4.355 | .000 | .070 | .186 |
| Wearing of PPE | .395 | 11.273 | .000 | .477 | .678 |

## 4. Discussion

Our study is novel as it explored how demographic and hygienic factors predict compliance of face mask wearing during a pandemic. Regardless of actions, the government has imposed to fight the Covid-19 pandemic. It is imperative to understand the roles of demographic and hygienic factors for at least two reasons. First, the compliance of citizens largely determines the effectiveness of a public health policy [18]. Second, understanding predictors would assist public officials to make informed decisions.

Our findings are consistent with recent studies on gender difference in terms of face mask wearing. A US study (n = 9,935) found that female shoppers were 1.5 times more likely to wear a face mask than male shoppers [19]. In a Saudi Arabia study (n = 1,767), women also showed better practice than men towards COVID-19 [20]. One possible explication would be that men might see face mask as infringing upon their independence, whereas women might only perceive face mask as being uncomfortable and be more willing to wear due to their self-protective instinct [21, 22]. In addition, women have a stronger tendency to feel shame from deviating from the norm and are more influenced by moral limitations [23], so they might be more compliant to face mask wearing.

Our results reveal that handwashing and use of PPE predict face mask wearing compliance. Both were necessary infection control measures during the SARS pandemic [24]. A SARS study found that handwashing alone reduced transmission by 55%, wearing PPE (gloves and gowns) by 57%, and wearing face mask by 68%; the cumulative effect of handwashing, face mask wearing and PPE reduced transmission by 91% [25]. The cumulative effect of precautions is the likely reason that handwashing and PPE are significantly related to face mask wearing.

This research is not without limitations. To begin with, our survey was conducted at a single point near the end of the first wave of the pandemic. It would be ideal for learning how people’s answers change as the pandemic enters the second or third wave. In terms of methodology, the web-based design means that people with no internet access and limited computer literacy were not surveyed, which explains the low percentage of respondents aged 60 or above in our sample (n = 55). Perhaps, the telephone survey should be complemented. Last but not least, other non-pharmaceutical interventions, such as physical distancing, are yet to be studied.

## 5. Conclusion

This study has identified several predictors of face mask wearing among adults during a pandemic, namely, gender, handwashing and PPE wearing. Gaining such understanding will assist public health organisations to come up with practical and targeted education campaign, and enforcement where appropriate. Future research may investigate predictors for face mask wearing compliance in a different country and cultural setting.

## Data Availability

Data are available upon official request.

## Notes

### Competing Interest Statement

The authors have declared no competing interest.

### Funding Statement

The authors and their institutions at any time received no payment or services from a third party for any aspect of the submitted work.

### Author Declarations

Full name of ethics committee / Institutional Review Board that ruled on ethics for this study is Xiamen University Malaysia Research Ethics Committee. Reference number is REC-2004.01

